# Evidence of Differences in Diurnal Electrodermal, Temperature, and Heart Rate Patterns by Mental Health Status in Free-Living Data

**DOI:** 10.1101/2024.08.22.24312398

**Authors:** Daniel McDuff, Isaac Galatzer-Levy, Seamus Thomson, Andrew Barakat, Conor Heneghan, Samy Abdel-Ghaffar, Jake Sunshine, Ming Zher-Poh, Lindsey Sunden, John Hernandez, Allen Jiang, Xin Liu, Ari Winbush, Benjamin W. Nelson, Nicholas B. Allen

## Abstract

Electrodermal activity (EDA) is a measure of sympathetic arousal that has been linked to depression in laboratory experiments. However, the inability to measure EDA passively over time and in the real-world has limited conclusions that can be drawn about EDA as an indicator of mental health status outside of controlled settings. Recent smartwatches have begun to incorporate wrist-worn continuous EDA sensors that enable longitudinal measurement of sympathetic arousal in every-day life. This work (N=237, 4-week observation period) examines the association between passively collected, diurnal variations in EDA and symptoms of depression, anxiety and perceived stress in a large community sample. Subjects who exhibited elevated depressive and anxiety symptoms had higher tonic EDA, skin temperature, and heart rate, despite not engaging in greater physical activity, compared to those that were not depressed or anxious. In contrast, subjects who exhibited elevated stress only exhibited higher skin temperature. Most strikingly, differences in EDA between those with high versus low symptoms were most prominent during the early morning. We did not observe amplitude or phase differences in the diurnal patterns. Our work suggests that electrodermal sensors may be practical and useful in measuring the physiological correlates of mental health symptoms in free-living contexts and that recent consumer smartwatches might be a tool for doing so.

## Introduction

Electrodermal activity (EDA) is a measure of sympathetic arousal, and has been widely adopted in psychophysiological research (Boucsein 2012). Tonic changes in the EDA signal - represented by the Skin Conductance Level (SCL) - have been reported as a useful feature for evaluating sympathetic arousal (Parsons et al. 2020; Kreibig 2010; Jacobs et al. 1994). However, the measurement of EDA in free-living contexts has been a longstanding challenge due to the paucity of convenient, unobtrusive and comfortable wearable measurement solutions. However, recent advances in wearable technology have led to the development of devices that address these limitations. For example, the Empatica E4^1^, Fitbit Sense 2^2^, and Google Pixel Watch 2^3^ all include dry electrode continuous EDA sensors integrated into their respective wrist-worn form factors. These devices continuously measure EDA throughout the day, capturing the full scope of daily arousals that can occur beyond the laboratory setting.

There are several studies that report differences in EDA in people experiencing depression (Iacono et al. 1983; Kim et al. 2018; Pruneti, Fiduccia, and Guidotti 2023). Monitoring EDA may help to differentiate the phases of depression (Sarchiapone et al. 2018) and anxiety (Birket-Smith, Hasle, and Jensen 1993); however, the limited studies in free-living settings have constrained the conclusions that can be drawn (Sarchiapone et al. 2018). The extant literature is based on laboratory studies that often analyze responses to contrived stimuli (Williams, Iacono, and Remick 1985). In-situ observations of diurnal variations in EDA are extremely limited, let alone such data for subgroups with psychiatric conditions.

Differences in electrodermal responses vary across psychopathologies. Electrodermal hyporeactivity (low variability) seems to be a reliable correlate of depressive symptoms (Sarchiapone et al. 2018) and has the ability to separate panic or agoraphobic groups from controls; however, participants with generalized anxiety disorder have shown little or no difference in EDA in some studies (Birket-Smith, Hasle, and Jensen 1993). Pruneti et al. (2023) evaluated changes in EDA in 53 patients with mixed anxiety and depressive disorders, and their results confirmed depression as the first predictor of suicidal thoughts but that EDA responses were a moderating factor such that those with high EDA showed a stronger association between depressive symptoms and suicidal thoughts.

While ambulatory, longitudinal SCL measurements are rarer in the literature, heart rate (Rykov et al. 2021) and skin temperature (Mason et al. 2024) have been reported on more systematically. Skin temperature, which is correlated with electrodermal activity (Khan et al. 2019), has also been observed to be higher in subjects with depression (Mason et al. 2024). We include analysis of these ancillary peripheral physiological measures of sympathetic ANS arousal to provide more context to the SCL findings.

Overall, in-situ/free-living monitoring of EDA in larger populations is essential to further our understanding of the relationships between EDA, depression, and anxiety in naturalistic settings. The introduction of commercial wearable devices equipped with continuous EDA sensors presents a considerable opportunity to both evaluate EDA in real-world contexts and to collect data on a scale that is sufficient to quantify meaningful differences. This represents a significant advancement in the field of psychophysiological research and has the potential to lead to new insights into the role of EDA in the diagnosis, monitoring, and treatment of mental health conditions. Recent research reported diurnal patterns in EDA based on observations of over 15,000 people who wore the Fitbit Sense 2, a commercially available smartwatch with an EDA sensor that continuously measures impedance on the dorsal wrist (McDuff, Thomson, et al. 2024).

In the current study, we present the in-situ evidence of deviations in diurnal EDA patterns that correlate with depression, anxiety, and perceived stress. Based on the previous literature, we hypothesized that elevated mental health symptoms would be associated with higher levels of EDA, and that diurnal patterns would reveal that this difference is strongest in the early morning. Finally, we compare these diurnal patterns with those from the large general population sample in our previous research (McDuff, Thomson, et al. 2024).

## Methods

### Study Design

The data for this study were collected as part of a prospective, nonrandomized study to investigate patterns and relationships between digital device use patterns, including sensor data from phones and wearables reflecting both behavioral and physiological processes, and self-reported measures of mental health and well-being. The study protocol (McDuff, Barakat, et al. 2024) was four weeks long with passive sensing from a wearable (Fitbit Sense 2), for the complete four-week period. The study was advertised via notifications in the Fitbit mobile application (Android version only). Any US resident adults aged 18 - 80 years old, who own an Android smartphone as their primary personal smartphone were eligible. The inability to provide informed consent via text-based instructions in English and major health conditions that severely restrict mobility and physical activity were the only exclusion criteria.

The data were collected with informed consent. After choosing to enroll, participants were directed to a consent form that described the data that were to be collected and how it would be used to advance the goals of the study. The data were de-identified and all 18 HIPAA identifiers (name, email, address, etc.) were removed from the study data. Passive data collection began only after participants signed a detailed informed consent and completed all onboarding surveys.

Data were collected in a manner that inherently restricts the granularity and detail of participant information. The electrodermal sensor was only activated on request by the participant (i.e., not on by default) and could be turned off at any point during the study without compromising their enrollment. In accordance with Fitbit Privacy Policy^3^, during the study participants could choose to delete their account, in which case their data would be deleted within 30 days.

At the start of the four-week study period, the participants completed a set of surveys which included the Patient Health Questionnaire (PHQ-8) (Kroenke et al. 2009), Generalized Anxiety Disorder Scale (GAD-7) (Löwe et al. 2008) and Perceived Stress Scale (PSS) (Cohen, Kamarck, and Mermelstein 1983).

### Participants

We recruited 395 participants who had a Fitbit Sense 2^TM^ device with the electrodermal sensor activated. Of these, 237 (female: 160, male: 68, queer: 6, other: 3) completed demographic, PHQ, GAD, and PSS intake surveys. The mean age was 45.1 years (st. dev. = 11.8 years). See Table 1 for the demographics. Participants exhibited a range of PHQ-8, GAD-7, and PSS scores (see Figure 1) that is consistent with other studies of broad populations (Sequeira et al. 2021). As expected there are covariations in these measures with the depressed group also more likely to be stressed and anxious (see Table 2). As such it is difficult to fully disentangle which traits drive the differences in behavior and physiology that we observe. Phenotypical SCL patterns may be due to a non-specific underlying mechanism common across depression, anxiety, and stress as there are highly comorbid conditions

**Table 1.** Demographics and Characteristics of Study Population.

| Factor (Level) | N = 237 | Percentage |
| --- | --- | --- |
| **Age (years)** | $\mu=45.1$ ($\sigma=11.8$) | |
| **Gender** |  |  |
| Female | 68 | 67.8% |
| Male | 160 | 28.4% |
| Queer/NonConforming | 6 | 2.5% |
| Other | 3 | 1.3% |
| **Sexual Orientation** |  |  |
| Heterosexual | 197 | 83.1% |
| LGBTQIA+ | 36 | 15.3% |
| I Prefer Something Else | 4 | 1.7% |
| **Race** |  |  |
| Caucasian | 207 | 87.7% |
| Black/African American | 8 | 3.4% |
| Asian/Pacific Islander | 3 | 1.3% |
| American Indian/Alaskan Native | 4 | 1.7% |
| Mixed Race | 10 | 4.2% |
| Other | 5 | 1.7% |
| **Ethnicity** |  |  |
| Hispanic or Latino | 220 | 92.8% |
| Not hispanic or Latino | 17 | 7.2% |
| **Characteristics** |  |  |
| Mild, Moderate, Severe Depression | 105 | 44.3% |
| Mild, Moderate, Severe Anxiety | 118 | 49.8% |
| High Stress | 143 | 60.3% |

<sup>3</sup> https://www.fitbit.com/global/us/legal/privacy-policy

**Table 2.** Cross Tabulations for PHQ-8 (>= 5 mild to high depression), GAD-7 (>= 5 mild to high anxiety) and PSS scores (>=14 high stress)

|  | Mild<br>Depress | Not<br>Depress |  | High<br>Stress | Not<br>Stress |  | Mild<br>Depress | Not<br>Depress |
| --- | --- | --- | --- | --- | --- | --- | --- | --- |
| **Mild-High**<br>**Anxiety** | 85 (36%) | 33 (14%) | **Mild-High**<br>**Anxiety** | 102 (43%) | 16 (7%) | **High**<br>**Stress** | 85 (36%) | 58 (24%) |
| **Not**<br>**Anxiety** | 20 (8%) | 99 (42%) | **Not**<br>**Anxiety** | 41 (17%) | 78 (33%) | **Not**<br>**Stress** | 20 (8%) | 74 (31%) |

**Table 3.** Non-linear function (CircaCompare [(Parsons et al. 2020)]) outputs when comparing not depressed (PHQ-8 <5) and mildly depressed or depressed groups (PHQ-8 >=5), not anxious (GAD-7 <5) and mildly anxious or anxious groups (GAD-7 >=5), and low stress (PSS<14) and moderate to high stress (PSS>=14) groups.

| Score | Param. | SCL |  | Skin Temp. |  | Heart Rate |  | Steps |  |
| --- | --- | --- | --- | --- | --- | --- | --- | --- | --- |
|  |  | Est. | 95% CI | Est. | 95% CI | Est. | 95% CI | Est. | 95% CI |
| PHQ-8 | Diff. in **Mesor** (k1) | 0.73* | [0.11 - 1.36] | 0.12* | [-0.01 - 0.25] | 2.81* | [0.63 - 4.99] | -0.10 | [-2.20 - 2.00] |
|  | Diff. in **Amplitude** (alpha1) | 0.14 | [-0.72 - 0.99] | -0.11 | [-0.26 - 0.03] | -0.40 | [-3.77 - 2.96] | -0.06 | [-2.77 - 2.66] |
|  | Diff. in **Phase** (psi1) | 2.08 | [-6.00 - 10.2] | -0.05 | [-0.50 - 0.40] | 0.02 | [-0.97 - 1.00] | 0.08 | [-1.32 - 1.48] |
| GAD-7 | Diff. in **Mesor** (k1) | 0.70* | [0.08 - 1.32] | 0.20* | [0.07 - 0.33] | 2.41* | [0.26 - 4.56] | 0.13 | [-1.97 - 2.24] |
|  | Diff. in **Amplitude** (alpha1) | 0.07 | [-0.79 - 0.93] | -0.05 | [-0.20 - 0.09] | -0.34 | [-3.56 - 2.88] | 0.02 | [-2.69 - 2.72] |
|  | Diff. in **Phase** (psi1) | 1.58 | [-6.32 - 9.49] | -0.17 | [-0.60 - 0.25] | -0.21 | [-1.20 - 0.77] | 0.08 | [-1.32 - 1.48] |
| PSS | Diff. in **Mesor** (k1) | 0.53 | [-0.07 - 1.13] | 0.09† | [-0.04 - 0.22] | 1.01 | [-1.30 - 3.32] | 1.00 | [-1.14 - 3.13] |
|  | Diff. in **Amplitude** (alpha1) | 0.03 | [-0.80 - 0.86] | 0.04 | [-0.11 - 0.19] | 0.25 | [-3.28 - 3.78] | 0.38 | [-2.39 - 3.15] |
|  | Diff. in **Phase** (psi1) | 1.33 | [-8.03 - 10.7] | -0.17 | [-0.61 - 0.27] | -0.17 | [-1.26 - 0.91] | -0.16 | [-1.62 - 1.29] |

\* = $p < 0.05$ , † = $p < 0.1$

**Table 4.** Fixed Effect Estimates, standard errors and estimated p-values from the Linear Mixed Effects Model.

| Predictor Variable | PHQ-8 | GAD-7 | PSS |
| --- | --- | --- | --- |
| *Intercept* | 3.40 | 4.51 | 13.9 |
| **Demographics** |  |  |  |
| Age | **-0.85\*** | **-1.18\*** | **-1.48\*** |
| Weight | **0.49\*** | 0.03 | **-0.48\*** |
| Gender (Female) | **1.57\*** | **0.94\*** | **1.88\*** |
| Orientation (LGBTQI+) | **3.43\*** | **1.08\*** | **2.39\*** |
| Race (Not Caucasian) | **-0.81\*** | **-0.93\*** | **-0.44\*** |
| **Phys./Behavior** |  |  |  |
| Mean Tonic SCL | **0.23\*** | 0.04 | **0.25\*** |
| Mean HR | -0.01 | **-0.06\*** | **0.08\*** |
| Mean Skin Temp. | **0.09\*** | **0.19\*** | **0.21\*** |
| Mean Steps | **-0.09\*** | **0.12** | 0.00 |

\* = $p < 0.01$

**Figure 1.**
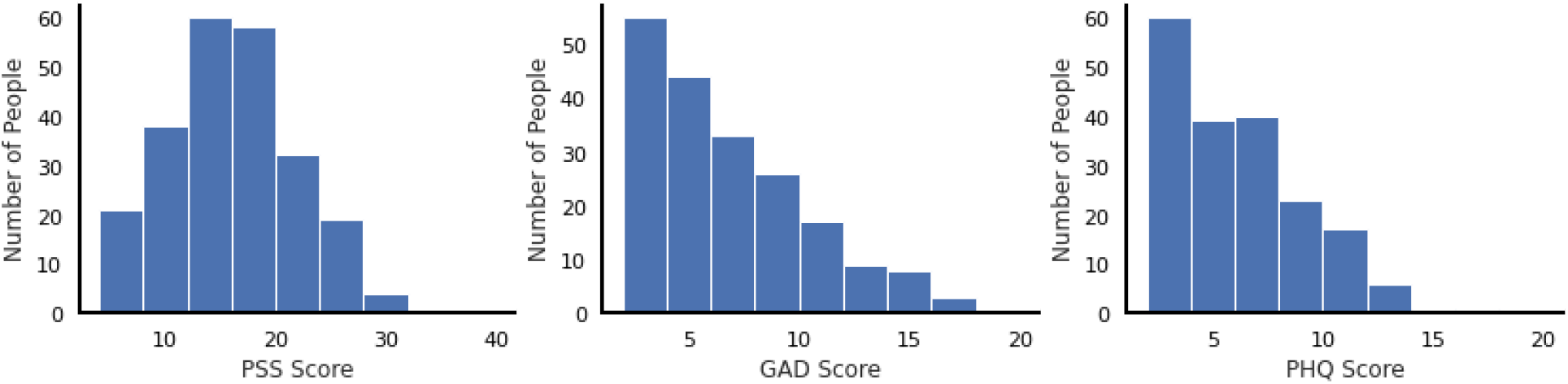
Distributions of PSS, GAD and PHQ scores in the population.

For additional context, we also compare observations from this study group to EDA data from a large general population sample of 15,349 US-based Sense 2 wearers (Men: 5,300, Women: 10,049; Mean Age: 49.8 years, Min: 18 years, Max: 91 years) for whom survey data is not available. Further EDA results from the large general population sample have been previously reported (McDuff et al., 2024).

### Sensor

Analyses used large-scale continuous EDA data captured on the commercially available smartwatch, Fitbit Sense 2, which is enabled with a dorsal wrist-facing EDA sensor. This sensor is positioned on the underside of the device wherein it is designed to directly interface with the skin and allow a continuous measurement of the dorsal wrist impedance. This impedance signal is acquired at 200 samples-per-second, which is then passed through a boxcar filter and down-sampled to 25 samples-per-second. This signal is then shifted and scaled using a benchtop calibration mapping that compares measured known impedance values to the internal electrical impedances of the device, from which admittance is calculated. For this study, the sensor data pulled from the device for analysis was median-filtered and then low-pass filtered minutely admittance values expressed in Siemens. Given the challenges of measuring EDA in ambulatory settings from the dorsal wrist we focused on tonic, rather than phasic, information. The EDA measurements were averaged at the minute-level thus removing higher frequency changes. In addition to EDA, the Fitbit Sense 2 is equipped with multiple sensing modalities including: multi-path optical heart rate tracker, ambient light sensor, gyroscope, 3-axis accelerometer, on-wrist skin temperature sensor, and red and infrared sensors for oxygen saturation monitoring.

We filter the signals to remove spurious measurements. The SCL values were filtered between 0 and 30 microsiemens, the HR between 40 and 250 beats/min, skin temperatures and steps were not filtered. The values were then aggregated by participant and hour to generate daily patterns from midnight to midnight. The resulting data have a positively skewed distribution. To preserve battery power the SCL sensor was off while participants were detected to be sleeping (the sensor was deactivated 15 minutes after sleep onset and reactivated 15 minutes after sleep offset), furthermore many users charge their device during the night or choose to sleep with it off.

Figure 2(a) shows the number of user/minutes with sensor data present for each minute in the day.

**Figure 2.**
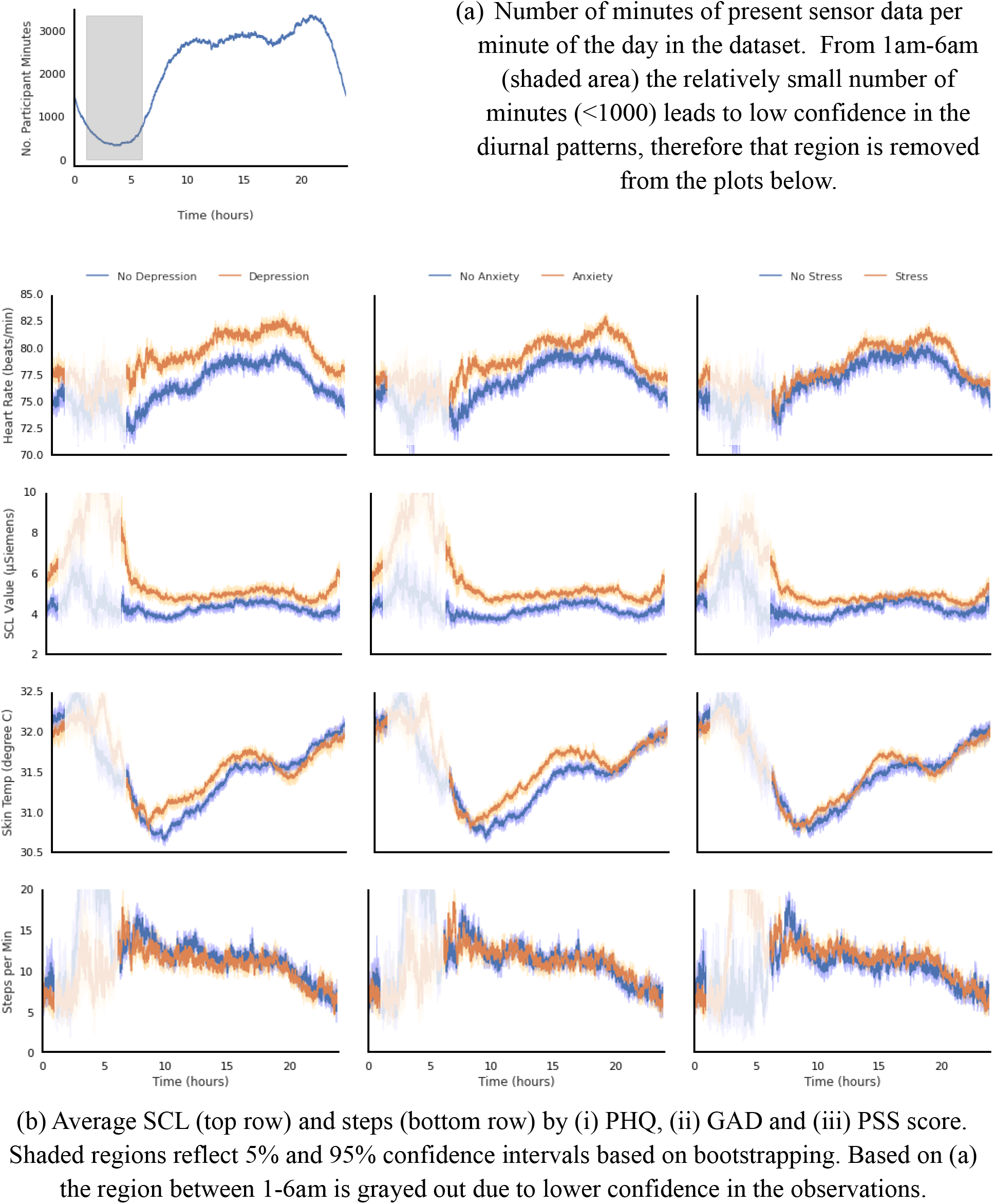
Diurnal patterns in physiological and behavioral measures across the dataset. The electrodermal sensor on the Fitbit Sense 2 was automatically turned off when the device detected the subject was sleeping. This contributes to larger uncertainty in measurement during typical sleeping hours (i.e., 10-6am).

### Model

We use a non-linear cosinor fitting method to estimate the difference in mesor, amplitude and phase, between the diurnal rhythms in HR, EDA, skin temperature and steps (CircaCompare [(Parsons et al. 2020)]). The form of the standard cosinusoidal curve is as follows with samples averaged per-user, per-hour:

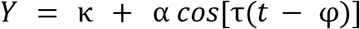

κ represents the mesor, a rhythm-adjusted mean, α represents amplitude, τ the period, refers to the time in hours and φ is the phase. The base equation was further modified to allow for the comparison between two sets of time series. The model tests for differences in mesor, amplitude and phase between two rhythms.

The CircaCompare package does not handle additional covariates, therefore to investigate whether the differences in mesors were a result of differences in population demographics we used a linear model fit via ordinary least squares regression to examine the strength of the effect between survey scores and tonic SCL, HR, skin temperature and steps while controlling for demographics. In this model we input the average SCL, HR, skin temperature and steps values per day and dummy code the gender, sexual orientation, race and ethnicity variables. For PHQ the following equation describes the model:

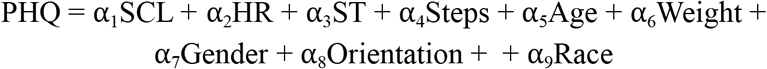

SCL, HR, ST, steps, age and weight were Z-scored and gender, orientation and race were dummy coded.

## Results and Discussion

In our cohort of 237 subjects, we observe elevated SCL levels in the depression (vs low depression) group. Figure 2 shows average HR, SCL, skin temperature and step counts as a function of the depression, anxiety and stress groupings. The plots illustrate that SCL is elevated, particularly in the depressed group and most prominently in the early morning/night time hours. This is supported when modeling the diurnal patterns using a cosinor model and also in an OLS model testing tonic SCL and controlling for demographics. Based on Figure 2 differences appear greatest from 6 am to noon; however due to the smaller number of samples during the 6-7am period there is also larger variance. Differences in SCL during the night-time/early morning hours between people with no depressive/anxiety symptoms and elevated depressive/anxiety symptoms deserve further research as our results suggest there might be phenotypical differences. Elevated tonic SCL in the depressed group is observed despite their taking no more steps, suggesting that the increased sympathetic arousal may be driven by factors other than physical activity. For context, Figure 3 shows a comparison between the groups in our study and baseline diurnal pattern from a sample of 15,349 Fitbit Sense 2 users (for whom there is no survey data) reported in prior work (McDuff, Thomson, et al. 2024). The control group (no depression, no anxiety, low stress) diurnal data in our study resemble the population level traces closely, particularly for SCL and heart rate measurement. Our results do not support theories that electrodermal hyporeactivity is a feature of depression in all contexts.

**Figure 3.**
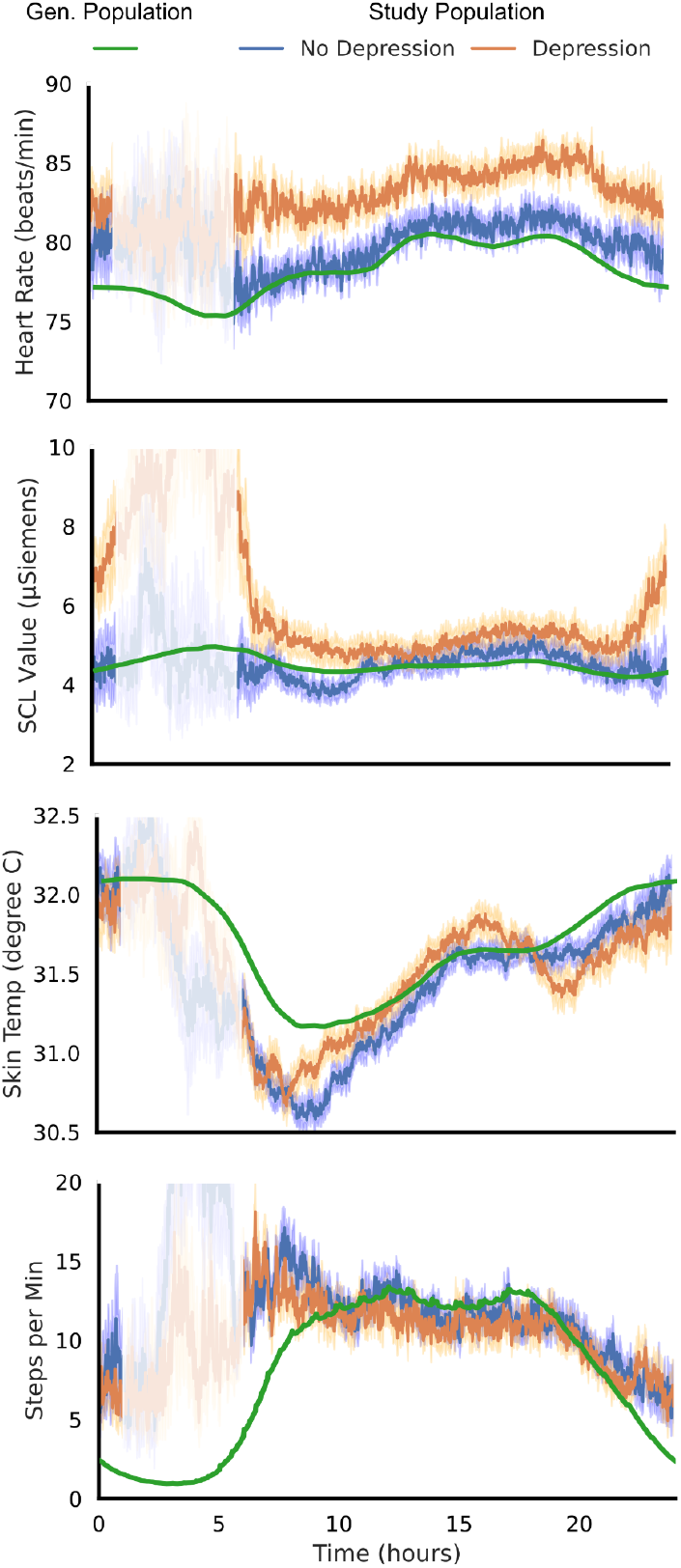
Comparison between study diurnal patterns (N=237) and population diurnal patterns (N=15,349)

Differences in physiological diurnal patterns were observed when comparing PHQ score, GAD score and PSS score in our cohort of 237 subjects. As expected, and consistent with high rates of comorbidity in the literature, in this population there is a considerable overlap in subjects who were depressed and anxious in particular (see covariance matrix in Table 2) and therefore similarity in the diurnal patterns is unsurprising. However, tonic SCLwas not significantly greater for the anxious group, consistent with prior work analyzing responses in lab conditions (Birket-Smith, Hasle, and Jensen 1993).

When we consider the other physiological variables we see patterns that confirm results in prior work. Skin temperature was slightly elevated in the depressed group, consistent with a recent large study in ambulatory settings of 20,000 subjects (Mason et al. 2024). The result was consistent in the cosinor model and the OLS model; however, the effects were smaller than for SCL. Heart rate was higher in the depressed group in the cosinor model; however, the tonic effect was no longer significant when controlling for other variables in the OLS model.

Longitudinal electrodermal sensing at scale has been made possible thanks to the miniaturization of electronics and advances in battery technology. It presents new opportunities for mental health features. Understanding normative patterns within the population and how these vary by mental health status is an important step in enabling more personalized and specific uses of these data.

Ultimately, the results demonstrate through the use of large scale sampling of physiological activity across a 24-hour period, that those with significant mood and anxiety disorder symptomatology display distinct patterns in their early morning physiological profile, particularly as expressed in electrodermal activity. This abnormal pattern is likely not mechanistic of mood and anxiety disorders, but rather likely reflects long-observed abnormalities in autonomic activity that have been observed in many downstream indicators. Examples include abnormalities in morning cortisol and other hormones and metabolites associated with hypothalamic-pituitary (HPA) axis and sympathetic-adrenal-medullary (SAM) system (Jacobson 2014). What is unique in the current work is a population profile of the physiology associated with autonomic activity as a consistent and discernible signal of those abnormal rhythms, making them measurable and actionable for predictive decision making at scale.

### Limitations

Despite various strengths including a month of continuous SCL, heart rate, skin temperature, and step data in a large sample using a novel smartwatch device, there are a number of limitations that should be mentioned. Stress in particular may vary greatly, 2) fairly mild (ie not severe depression or anxiety - PHQ-8 cut off of 5 is pretty low) which could maybe mask differences you might see with more severe anxiety or depression, 3) comorbidity was high amongst participants with anxiety and depression.

Wear time of the sensors was marginally higher for subjects who were not depressed (32.0% of minutes during the 28 days) compared to those that had mild, moderate, moderately-severe, or severe depression (30.5% of minutes).

Measuring any physiological signals in ambulatory settings is non-trivial. There are several potential confounders/sources of noise. Participants who wear devices more loosely on their wrists will have less reliable measurements from all the sensors, including SCL. Measurements during exercise are less reliable than when subjects are stationary, while exercising happens only a minority of the time during the day it can have an outsized effect on higher heart rate and skin temperature measurements. The significant results we observed did not appear attributable to exercise effects. Finally, given that SCL sensors are measuring conductance, in the presence of water, such as when someone takes a shower, they can return very high values when the subject or the device gets wet.

## Conclusion

This study investigated whether longitudinally collected Fitbit Sense 2 smartwatch-derived SCL, heart rate, skin temperature, and step count differed between participants with elevated depressive, anxiety, and stress symptoms as compared to healthy controls across 24 hours. Overall, results indicate that participants with elevated depressive and anxiety symptoms had higher tonic EDA, skin temperature, and heart rate, despite not engaging in greater physical activity, compared to those who were not depressed or anxious. In contrast, participants with elevated stress only had higher skin temperature. The most prominent temporal difference between participants with clinically elevated symptoms and healthy controls were EDA measurements that occurred in the early morning. Our results suggest ambulatory SCL measured from the dorsal wrist can be practical and useful in measuring the severity of mental health symptoms in free-living contexts.

## Data Availability

We support open science principles, but also recognize that it is often challenging to ensure participant privacy while also making data broadly available to the academic research community. We had to balance these considerations with the privacy of the participants and protection of their health data. Furthermore, although the data could be de-identified, some of the data streams could not be fully anonymized. We recognize that this is a limitation, but we felt that we needed to give the participants strong reassurance that their data would not be used for any other purpose than for the research at hand.

## Supplementary Materials

**Table S1.** Non-linear function (CircaCompare) outputs when comparing not depressed (PHQ<5) and depressed groups (PHQ>=5)

| Param. | SCL |  | Skin Temp. |  | Heart Rate |  | Steps |  |
| --- | --- | --- | --- | --- | --- | --- | --- | --- |
|  | Est. | 95% CI | Est. | 95% CI | Est. | 95% CI | Est. | 95% CI |
| Mesor (k) | 2.23 | [1.85 - 2.60] | 31.5 | [31.4 - 31.6] | 76.4 | [75.0 - 77.9] | 6.33 | [4.91 - 7.75] |
| Diff. in Mesor (k1) | **0.73\*** | **[0.11 - 1.36]** | **0.12\*** | **[-0.01 - 0.25]** | **2.81\*** | **[0.63 - 4.99]** | -0.10 | [-2.20 - 2.00] |
| Amplitude (alpha) | 0.08 | [-0.35 - 0.50] | 0.51 | [0.41 - 0.61] | 2.92 | [0.66 - 5.18] | 2.14 | [0.37 - 3.91] |
| Diff. in Amplitude (alpha1) | 0.14 | [-0.72 - 0.99] | -0.11 | [-0.26 - 0.03] | -0.40 | [-3.77 - 2.96] | -0.06 | [-2.77 - 2.66] |
| Phase (psi) | 5.47 | [-2.09 - 13.0] | 5.57 | [5.32 - 5.83] | 4.06 | [3.44 - 4.67] | 3.01 | [2.06 - 3.96] |
| Diff. in Phase (psi1) | 2.08 | [-6.00 - 10.2] | -0.05 | [-0.50 - 0.40] | 0.02 | [-0.97 - 1.00] | 0.08 | [-1.32 - 1.48] |

\* = $p < 0.05$

**Table S2.**
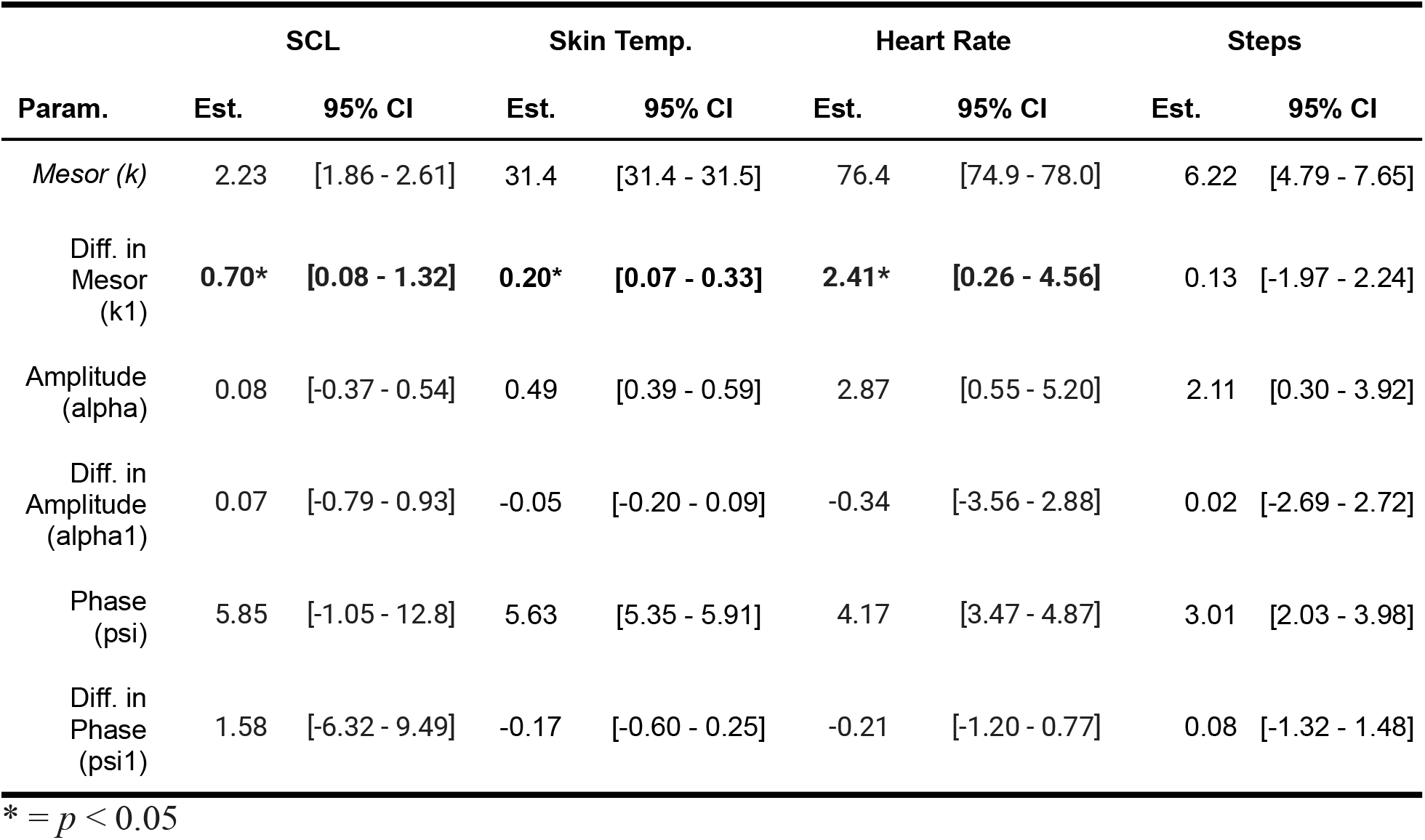
Non-linear function (CircaCompare) outputs when comparing not anxious (GAD<5) and anxious groups (GAD>=5)

| Param. | SCL |  | Skin Temp. |  | Heart Rate |  | Steps |  |
| --- | --- | --- | --- | --- | --- | --- | --- | --- |
|  | Est. | 95% CI | Est. | 95% CI | Est. | 95% CI | Est. | 95% CI |
| *Mesor (k)* | 2.23 | [1.86 - 2.61] | 31.4 | [31.4 - 31.5] | 76.4 | [74.9 - 78.0] | 6.22 | [4.79 - 7.65] |
| Diff. in Mesor (k1) | **0.70\*** | **[0.08 - 1.32]** | **0.20\*** | **[0.07 - 0.33]** | **2.41\*** | **[0.26 - 4.56]** | 0.13 | [-1.97 - 2.24] |
| Amplitude (alpha) | 0.08 | [-0.37 - 0.54] | 0.49 | [0.39 - 0.59] | 2.87 | [0.55 - 5.20] | 2.11 | [0.30 - 3.92] |
| Diff. in Amplitude (alpha1) | 0.07 | [-0.79 - 0.93] | -0.05 | [-0.20 - 0.09] | -0.34 | [-3.56 - 2.88] | 0.02 | [-2.69 - 2.72] |
| Phase (psi) | 5.85 | [-1.05 - 12.8] | 5.63 | [5.35 - 5.91] | 4.17 | [3.47 - 4.87] | 3.01 | [2.03 - 3.98] |
| Diff. in Phase (psi1) | 1.58 | [-6.32 - 9.49] | -0.17 | [-0.60 - 0.25] | -0.21 | [-1.20 - 0.77] | 0.08 | [-1.32 - 1.48] |

\* = $p < 0.05$

**Table S3.** Non-linear function (CircaCompare) outputs when comparing low stress (PSS<14) and moderate-high stress groups (PSS>=14)

| Param. | SCL |  | Skin Temp. |  | Heart Rate |  | Steps |  |
| --- | --- | --- | --- | --- | --- | --- | --- | --- |
|  | Est. | 95% CI | Est. | 95% CI | Est. | 95% CI | Est. | 95% CI |
| *Mesor (k)* | 2.22 | [1.78 - 2.66] | 31.5 | [31.4 - 31.6] | 77.1 | [75.3 - 78.9] | 5.69 | [4.06 - 7.32] |
| Diff. in Mesor (k1) | 0.53 | [-0.07 - 1.13] | 0.09<sup>†</sup> | [-0.04 - 0.22] | 1.01 | [-1.30 - 3.32] | 1.00 | [-1.14 - 3.13] |
| Amplitude (alpha) | 0.08 | [-0.47 - 0.62] | 0.44 | [0.32 - 0.56] | 2.53 | [-0.20 - 5.26] | 1.91 | [-0.21 - 4.03] |
| Diff. in Amplitude (alpha1) | 0.03 | [-0.80 - 0.86] | 0.04 | [-0.11 - 0.19] | 0.25 | [-3.28 - 3.78] | 0.38 | [-2.39 - 3.15] |
| Phase (psi) | 5.99 | [-2.25 - 14.2] | 5.65 | [5.30 - 6.01] | 4.19 | [3.30 - 5.07] | 3.16 | [1.97 - 4.34] |
| Diff. in Phase (psi1) | 1.33 | [-8.03 - 10.7] | -0.17 | [-0.61 - 0.27] | -0.17 | [-1.26 - 0.91] | -0.16 | [-1.62 - 1.29] |

\* = $p < 0.05$ , <sup>†</sup> = $p < 0.1$

## Footnotes

1 https://www.empatica.com/research/e4/

2 https://www.fitbit.com/global/us/products/smartwatches/sense2

3 https://www.fitbit.com/global/us/legal/privacy-policy

